# Assessing Willingness to receive COVID-19 Vaccines, associated factors and reasons for hesitancy among persons aged 13-80 years in Central Uganda. A population-based surveillance Cohort

**DOI:** 10.1101/2023.04.19.23288804

**Authors:** Alex Daama, Naziru Rashid, Kasango Asani, Grace Kigozi Nalwoga, Fred Nalugoda, Robert Bulamba, Emmanuel Kyasanku, Gertrude Nakigozi, Godfrey Kigozi, Joseph Kagaayi, Stephen Mugamba

**Author notes:** Corresponding author: Alex Daama. Africa Medical and Behavioral Sciences Organization (AMBSO). Plot 7441, Nansana, Hoima Road, Wakiso, Uganda P.O Box 25974 Wakiso. E-mail addresses of authors: AD.

## Abstract

**Background:** Vaccination is essential for controlling the COVID-19 pandemic. However adequate vaccine coverage is a critical to the effectiveness of the vaccine at a population level. Data on to acceptability of the vaccine in Urban areas are limited. This study examined the prevalence, factors associated with willingness to receive COVID-19 vaccine and reasons for hesitancy in the predominantly urban in central Uganda (Wakiso)

**Methods:** Data were obtained from a cross-sectional study conducted from March 1st, 2021, to September 30th, 2021 in the urban population-based cohort of the Africa Medical and behavioral Sciences Organization (AMBSO). Multivariable modified Poisson regression analysis was used to estimate adjusted prevalence ratios (aPR) and 95% confidence intervals of willingness to accept the COVID-19 vaccine.

**Results:** A total of 1,903 participants were enrolled in the study; 61% of whom were females. About 63% of participants indicated willingness to accept the COVID-19 vaccine. Younger age groups (13-19 and 20-29) were less likely to accept the vaccine compared to the persons ages 40-49 years (aPR=0.79; 95% CI: 0.74, 0.84 for the 13-19 years and 0.93; 95% CI: 0.88, 0.98 for age group 20-29, compared to those ages 40–49 years. Post-primary education (aPR=1.05; 95% CI: 1.02, 1.09 compared to primary level), being a students and government staff (APR=1.13; 95% CI: 1.04, 1.23 compared to construction and Mechanic workers) were associated with willingness to receive COVID-19 vaccine. Some of the reported reasons for hesitancy included; concerns about side effects 154(57.0%), about 64(23.7%) did not think the vaccines were effective, and those who did not like the vaccines 32(11.9%).

**Conclusion:** A substantial proportion of individuals were not willingness to receive the COVID-19 vaccine. More effort is needed to reduce vaccine hesitancy, especially among the young and people with lower formal education.

## Introduction

The Corona virus disease 2019 (Covid-19), is a respiratory illness caused by Novel Corona Virus also known as Severe Acute Respiratory Syndrome Corona Virus 2 (SARS Cov.2) (1). The disease is characterized by; dry cough and shortness of breath with difficulty in breathing and at least 2 of the following: fever, chills, muscle pain, headache, sore throat and loss of test and smell. The control of the disease is still through preventive measures including but not limited to hand hygiene (frequent hand washing and sanitization) keeping social distance, use of face masks and vaccination. Except vaccination, a population based Ugandan study reported a higher uptake of COVID-19 preventive measures (2).

Globally as of 31^st^ January 2023, there were about 753 million confirmed cases of Covid-19 with about 6.8 million deaths (3). Africa reported about 9.4 million confirmed cases and 175,247 deaths (3). Uganda has so far reported 170,233 confirmed cases of Covid-19 with 3,630 deaths as of 31^st^ January 2023 (3). These statistics show us that there was a great impact on the health care system of various countries. This therefore calls for tight measures in preventing and controlling Covid-19 thus the need of ensuring vaccination of the majority of members within the community.

Vaccination has been and is still very important in controlling communicable diseases including COVID-19. A number of COVID-19 vaccines have been developed and put on the market including AstraZeneca, Moderna, Pfizer, Johnson and John among others. For vaccination campaigns to be effective a critical mass of the community must be vaccinated to achieve a herd immunity (4). WHO Statistics show that as of January 2023 about 13 billion doses of doses of covid-19 have been administered globally. 28.21 % of the Africa’s population has received the last dose of Covid-19 vaccination and 28.34% of the Ugandan Population has received at least 2 doses of COVID 19 Vaccines (3).

These statistics show that uptake of COVID-19 vaccination and in Africa and in Uganda specifically is still low. Studies have found varying levels of willingness to up vaccination in various parts of the world. A study done among college students in Ethiopia showed that 34.2% of them were willing to take up COVID-19 vaccination. (5). A similar study done among people with preexisting health conditions in northwest Ethiopia found that 54.6% of this category of people were willing to take up COVID-19 vaccination (6). However a study done among elderly group of people in Canada found a higher prevalence of willingness to take up covid-19 vaccination (7). And finally, a study done at the Islamic university in Uganda found COVID 19 vaccination uptake at 20.4% (8).

Several factors have been highlighted regarding willingness to take up COVID 19 vaccination including gender, marital status, level of education, occupation exposure to COVID-19 and watching television (4,5). Being a health care worker, having lost someone, having good knowledge about vaccine perceived susceptibility to infection and the effectiveness of the vaccine have also been mentioned as factors affecting the willingness to take covid-19 vaccination (7,9). There are different reasons for why some people hesitate or refuse to receive COVID-19 vaccine as described elsewhere and these include; fear of side effects, vaccines not being effective, don’t like vaccines, COVID-19 is not a serious illness, Concern about the costs associated with the vaccine (such as office visit costs or vaccine administration fees), distrust in vaccines (10,11) adults (12). However, many of these studies have been conducted in high income countries and among students for example Spain, Malaysia with limited urban population-based studies in Sub-Saharan Africa more especially Uganda (Wakiso) among persons aged 13-80 years. Therefore, this study is timely to close the gap. Although, a Ugandan study was conducted to investigate reasons for unwillingness to participate in COVID-19 vaccine (13). This was basically a facility-based study and focused on health workers and hence limited data is available about reasons for hesitation in the general populations in the communities. In order to have a successful vaccination program to achieve herd immunity within the population, the factors determining the willingness to receive Covid-19 vaccination and reasons for hesitation must be well understood. The current study therefore looked at the prevalence and associated factors of willingness to receive COVID-19 vaccination including reasons for hesitancy in Central Uganda (Wakiso).

## Materials and Methods

### Study setting and design

The cross-sectional survey was conducted in urban settings of central Uganda (Wakiso) in the existing Population Health Surveillance (PHS) being implemented by Africa Medical and Behavioral Sciences Organization (AMBSO). PHS is an open urban population-based cohort which enrolls about 5,000 consenting adults 13-80 years in three communities of Wakiso and these include Kazo, Lukwanga and Sentema. The AMBSO-PHS aims have been described (14).

### Study population

The study population were all persons aged 13-80 years in Central Uganda (Wakiso).

### Inclusion criteria

A person was included in the study if him/her; was aged 13-80 years and consented to be interviewed. All members of the PHS cohort aged (13-80) years with non-missing data on willingness to receive COVID-19 vaccine were considered for analysis

### Exclusion criteria

Individuals with physical and mental impairment that interfere with their ability to offer informed consent and participate in this study.

### Sampling procedure

Firstly, census was conducted in the PHS selected areas that were done initially through community mapping exercise and compared population structure with distances from urban centres and cultivation. The overall objective was to select representative sample of households in the different community.

### Data collection

Census activities were done first which included household enumeration of eligible participants, obtaining information such as sex, age, relationship to head of household, marital status, and residence status. After census activities, all participants that were eligible were invited for a nearby venue called a hub where PHS survey/data collection take place and this was done for approximately 1-2 months per community for the annual surveillance as survey round three (R3). At the hub same sex interviews were conducted by experienced AMBSO data collectors and biological samples were drawn including anthropometrics measurements for vitals. Consent was obtained before any data collection activities. Data was collected on both paper and electronically.

Sample size: The sample size of this study was 1,903 participants based on the available captured data in the Population Health Surveillance in the three communities in surveillance visit three as per inclusion criteria

### Data analysis

We analyzed this data using the help of Stata version 15 (64-bit) software. Prevalence of willingness to receive COVID-19 vaccine and hesitation were determined through cross-tabulation. Descriptive and univariable analysis was performed to describe how the variables, reasons for hesitation or refusal as distributed amongst the participants. The frequency distributions for categorical variables were computed. For the continuous variables, we analyzed them preliminarily by use of means (with the standard deviation), modes and median (with the range). Bivariate analysis was conducted between each independent factor against willingness to receive COVID-19 status to determine relationships, respectively. Those variables known to be associated with smoking with a p-value < 0.2 as well as those which are biologically plausible were included in the multivariable model. Multivariate analysis was conducted to estimate odds ratios and 95% confidence intervals for the association between factors and willingness to receive COVID-19.

### Ethical considerations

Informed consent was obtained from all participants as per declaration of Helsinki ethical guidelines. The AMBSO-PHS protocol was reviewed and approved by Clark International University Research and Ethics Committee (UG-REC-015-CIURE/0059) and registered with the Uganda National Council for Science and Technology. Only de-identified participant and household IDs are used to link all participant data, which is pseudonymized throughout. The information is then maintained in secure locations or on computers that only authorized specific AMBSO data managers and editors have access to.

## Results

This study included 1,903 participants and the mean age of participants was 30.9 (SD=14.0) years (Table 2). Of the study participants, (1,170 [61.5%]) were female; (1,015 [53.4%]) were single.

**Table 1:**
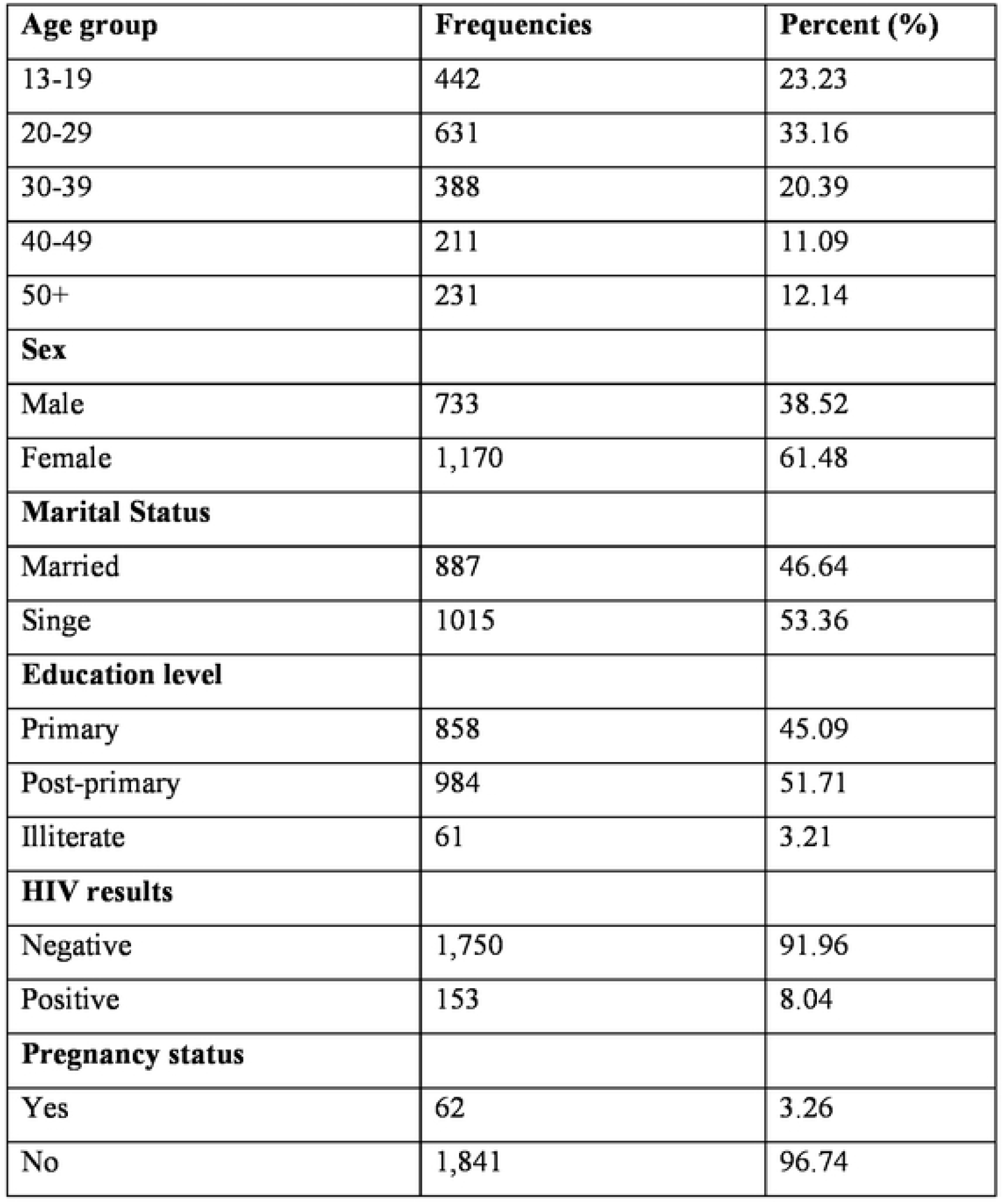
Showing socio-demographic characteristic of respondents.

| **Age group** | **Frequencies** | **Percent (%)** |
| --- | --- | --- |
| 13-19 | 442 | 23.23 |
| 20-29 | 631 | 33.16 |
| 30-39 | 388 | 20.39 |
| 40-49 | 211 | 11.09 |
| 50+ | 231 | 12.14 |
| **Sex** |  |  |
| Male | 733 | 38.52 |
| Female | 1,170 | 61.48 |
| **Marital Status** |  |  |
| Married | 887 | 46.64 |
| Singe | 1015 | 53.36 |
| **Education level** |  |  |
| Primary | 858 | 45.09 |
| Post-primary | 984 | 51.71 |
| Illiterate | 61 | 3.21 |
| **HIV results** |  |  |
| Negative | 1,750 | 91.96 |
| Positive | 153 | 8.04 |
| **Pregnancy status** |  |  |
| Yes | 62 | 3.26 |
| No | 1,841 | 96.74 |

**Table 2:** showing reasons for COVID-19 vaccine hesitancy among persons aged at least 13 years.

| **Reasons for not taking the Vaccine** | **Frequencies** | **Percent (%)** |
| --- | --- | --- |
| **Concern about side effects** |  |  |
| Yes | 154 | 57.04 |
| No | 116 | 42.96 |
| **Not worried about getting seriously ill from Corona virus infection** |  |  |
| Yes | 18 | 6.67 |
| No | 252 | 93.33 |
| **Don't think vaccines are effective** |  |  |
| Yes | 64 | 23.7 |
| No | 206 | 76.3 |
| **Don't like vaccines** |  |  |
| Yes | 32 | 11.85 |
| No | 238 | 88.15 |
| **Allergic to vaccines** |  |  |
| Yes | 02 | 0.74 |
| No | 268 | 99.26 |
| **Don't have time to get vaccinated** |  |  |
| Yes | 02 | 0.74 |
| No | 268 | 99.26 |
| **Concerns about getting corona virus infection from the vaccine** |  |  |
| Yes | 9 | 3.33 |
| No | 261 | 96.67 |
| **Other conspiracy theories mentioned** |  |  |
| Yes | 24 | 8.89 |
| No | 246 | 91.11 |
| **Was not eligible because had COVID 19 in the past 6 months, /pregnant, /breast feeding** |  |  |
| Yes | 4 | 1.48 |
| No | 266 | 98.52 |
| **Vaccine was not available, / distance to the facility was too long** |  |  |
| Yes | 2 | 0.74 |
| No | 268 | 99.26 |
| **Other reasons** |  |  |
| Yes | 22 | 8.15 |
| No | 249 | 91.85 |

Most participants (629 [57.6%]) had post-primary education while, (431 [39.5%]) had primary level, and only (32 [2.9%]) had no education. Majority of the participants were Christians (75 [71.0%]), Muslims (310 [28.4%]) and (7 [0.6%]) had no religious affiliation. Occupation groups included traders (318 [29.1%]); housework; (242 [22.2%]), agriculture (126 [11.5%]); construction & mechanics workers (120 [11.0%]), Boda-boda men (60 [5.49%]) and other had other occupations (175 [16.0%]). About (74 [6.8%]) of participants were HIV-positive and (753 [69.0%]) drank alcohol.

### Prevalence of willingness to receive COVID-19 vaccine

In this study, the prevalence of willingness to receive COVID-19 was 1206 (63.4%) while hesitation was 697(36.6%) as shown in fig 1.

**Figure 1:**
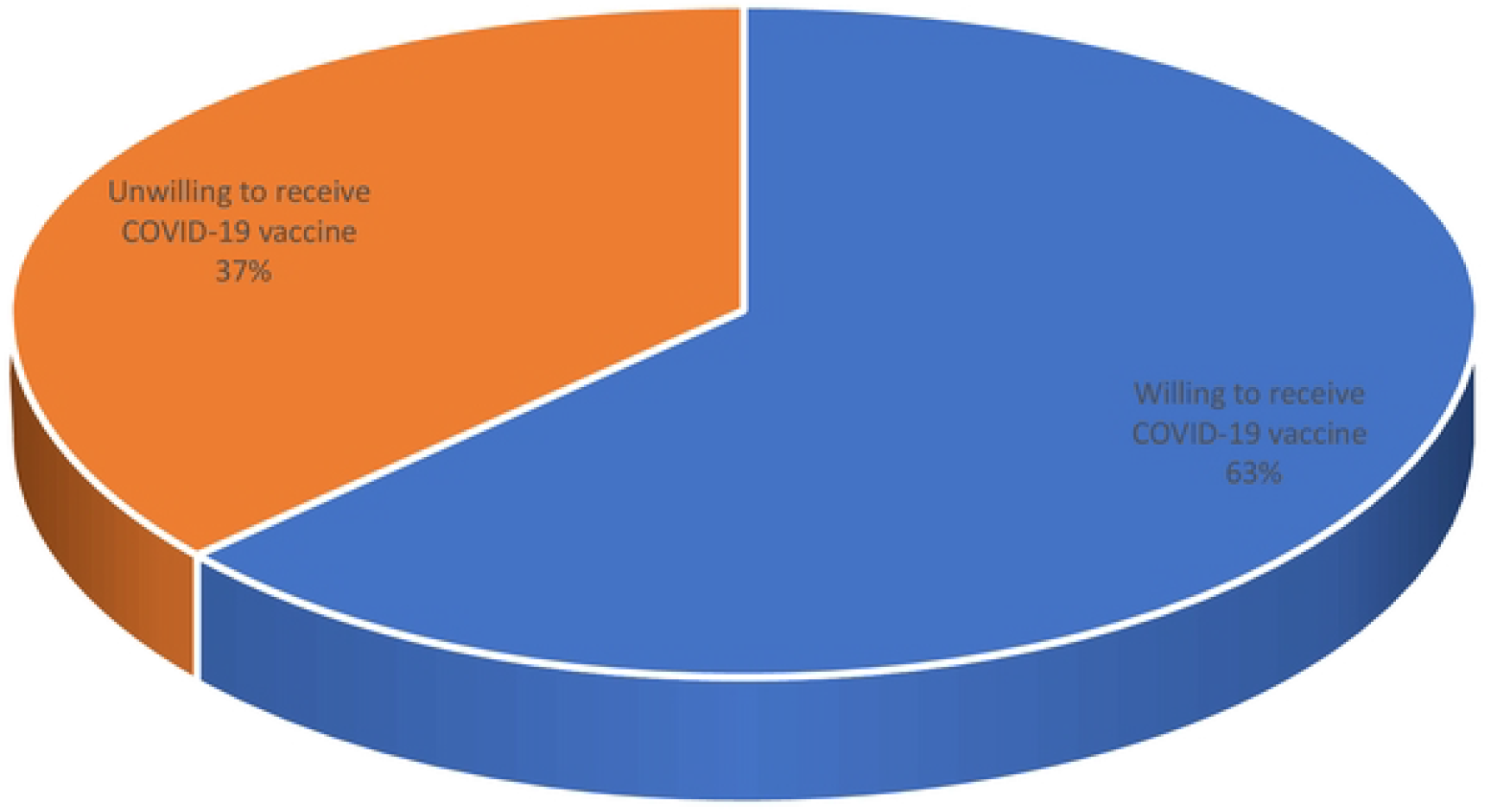
A piechart showing prevalence of willingness to taking COVID-19 Vaccine.

We investigated reasons for hesitation or refusal to receive COVID-19 vaccine among persons aged 13-80 years. The participants were asked different questions to assess reasons for unwillingness to receive COVID-19 vaccine (Table 2). Of the 11 reasons that were reported, majority of the participants had concerns about side effects resulting from the vaccines 154(57.0%), about 64(23.7%) did not think the vaccines were effective, this was followed by those who did not like the vaccines 32(11.9%). Besides that, about 24(8.9%) were unwilling because of other conspiracy theories. Additionally, other reasons included; no worries about getting seriously ill from the COVID-19 infection 18(6.7%), only 02(0.7%) reported that long distance to facility or vaccine not being available was the main reason for refusal or hesitancy.

In a bivariate analysis (Table 3), a variety of factors are associated with willingness to receive COVID-19 vaccine for example, older youths (25-29) and (30-35) years were more likely to smoke compared to those aged 18-24 years; (cOR=0.83 [95% CI: 0.78, 0.87], p<0.0001 (cOR=1.05, [95% CI: 21.02, 1.09], p=0.001) respectively. Other factors that had higher likelihood of smoking included alcohol use (cOR=1.11 [95% CI: 1.02, 1.22], p=0.016), HIV positive status (cOR=1.04, [95% CI: 1.00, 1.16], p=0.073)), (cOR=1.11 95% CI: 1.01, 1.22), p=0.027. Lastly, marital status, education, religion, and occupation were not associated with tobacco use.

**Table 3:** Showing factors associated with willingness to receive COVlD-19 vaccine.

| **Variables** | **Unadjusted**<br>**PR** | **P-value** | **[95% CI]** | **Adjusted**<br>**PR** | **P-value** | **[95% CI]** |
| --- | --- | --- | --- | --- | --- | --- |
| **Sex** |  |  |  |  |  |  |
| Male | 1.00 |  |  | 1.00 |  |  |
| Female | 1.03 | 0.087 | 0.99, 1.06 | 1.01 | 0.501 | 0.98, 1.05 |
| **Age group** |  |  |  |  |  |  |
| 40-49 | 1.00 |  |  | 1.00 |  |  |
| 13-19 | 0.83 | <0.0001 | 0.78, 0.87 | 0.79 | <0.0001 | 0.74, 0.84 |
| 20-29 | 0.95 | 0.066 | 0.90, 1.00 | 0.93 | 0.012 | 0.88, 0.98 |
| 30-39 | 0.97 | 0.285 | 0.91, 1.03 | 0.96 | 0.138 | 0.90, 1.01 |
| 50+ | 1.01 | 0.655 | 0.95, 1.08 | 1.02 | 0.457 | 0.96, 1.09 |
| **Educational level** |  |  |  |  |  |  |
| Primary | 1.00 |  |  | 1.00 |  |  |
| Post-primary level | 1.05 | 0.001 | 1.02, 1.09 | 1.05 | 0.002 | 1.02, 1.09 |
| Illiterate | 1.11 | 0.016 | 1.02, 1.22 | 1.08 | 0.097 | 0.99, 1.18 |
| **Chronic diseases** |  |  |  |  |  |  |
| No | 1.00 |  |  | 1.00 |  |  |
| Yes | 1.04 | 0.073 | 0.99, 1.09 | 0.98 | 0.42 | 0.93, 1.03 |
| **Occupations** |  |  |  |  |  |  |
| Construction and Mechanic workers | 1.00 |  |  | 1.00 |  |  |
| Agriculturalists | 1.03 | 0.403 | 0.96, 1.11 | 1.04 | 0.294 | 0.96, 1.13 |
| Housework | 1.08 | 0.056 | 1.00, 1.16 | 1.05 | 0.278 | 0.97, 1.13 |
| Traders/Vendors | 1.05 | 0.247 | 0.97, 1.13 | 1.01 | 0.83 | 0.93, 1.09 |
| Students and Govt staff | 1.04 | 0.336 | 0.96, 1.12 | 1.13 | 0.005 | 1.04, 1.23 |
| Other occupations | 1.11 | 0.027 | 1.01, 1.22 | 1.09 | 0.076 | 0.99, 1.20 |

In the multivariable analysis, as per the set criteria, there are variables at bivariate analysis that were not included in the final model including religion, occupation, and marital status.

In a multivariable model, younger age groups (13-19 and 20-29) were less likely to accept the vaccine compared to the persons ages 40-49 years (aPR=0.79; 95% CI: 0.74, 0.84 for the 13-19 years and 0.93; 95% CI: 0.88, 0.98 for age group 20-29, compared to those ages 40–49 years. Post-primary education (aPR=1.05; 95% CI: 1.02, 1.09 compared to primary level), being a students and government staff (APR=1.13; 95% CI: 1.04, 1.23 compared to construction and Mechanic workers) were associated with willingness to receive COVID-19 vaccine.

## DISCUSSION

About 63% of participants indicated willingness to accept the COVID-19 vaccine. This prevalence is slightly higher than the prevalence of 57% that was reported in South Africa (15). Another study done in Ethiopia reported that 54.6% of the participants were willing to receive the COVID-19 vaccine (9). Low willingness to accept vaccination were reported in Nigeria and Ethiopia (46.2%, and 33.7%) respectively (9). Another study done in Ethiopia that assessed willingness to receive COVID-19 vaccine among college students in Gondor city reported that only 32% of the participants were willing to receive the vaccine (5). Furthermore, a study done among residents of South Western Ethiopia reported that only 29% of the participants were willing to receive the vaccine (19). The relatively high willingness to accept the vaccine could be attributed to the vigorous campaigns by the government of Uganda to promote COVID-19 vaccination (20). Conversely, this prevalence is slightly lower than a prevalence of 72.7% which was reported in Kampala Uganda by (21). Another study done in Ireland reported that 72% of their participants were willing to receive COVID-19 vaccine (22). A study done in Ghana reported that 70% of the participants were willing to receive COVID-19 vaccine (23). A study done among undergraduate students in Southern Nigeria reported that 73% of the students were willing to receive COVID-19 vaccine (24). Another study done in Canada reported that 84% of the participants were willing to vaccinate (7). The relatively lower prevalence of willingness to receive COVID-19 vaccine in our study could be attributed to the differences in methodologies and the sociodemographic characteristics of population studied.

We examined some of the reasons why there is hesitation or refusal to receive COVID-19 vaccine among persons aged 13-80 years and majority of the participants had concerns about side effects resulting from the vaccines. These findings are consistent by a study conducted in Sentinel Schools Network of Catalonia, Spain conducted among students and parents (10). Besides our findings also revealed that vaccines were not liked and that these vaccines could infect them with the Corona virus. We found similar findings from Malaysia, as a study conducted by Kai Wei Lee et al among students (11). Furthermore, our findings indicated that some people were hesitant because they had not COVID 19 in the past 6 months, /pregnant, /breast feeding and therefore ineligible for vaccine uptake. This finding agrees with a massive online survey conducted among US adults (12). These used similar methodologies which were the ideal methods based on the outcome.

Younger age groups (13-19 and 20-29) were less likely to accept the vaccine compared to the persons ages 40-49 years (aPR=0.79; 95% CI: 0.74, 0.84 for the 13-19 years and 0.93; 95% CI: 0.88, 0.98 for age group 20-29, compared to those ages 40–49 years. This finding is consistent with findings from studies done in Africa which also reported that young participants were less likely to accept COVID-19 vaccine compared to old participants (19,25,26). The possible reason is because younger people do not perceive themselves to be at risk of developing COVID related complications as compared to the ageing population (26,27). Post-primary education (aPR=1.05; 95% CI: 1.02, 1.09 compared to primary level). This is consistent with findings from (5,19,28,29). The possible explanation could be that higher education level increases the level of knowledge and awareness about COVID-19 and its prevention strategies; including vaccination and therefore willingness to vaccinate. Students and government staff were also willing to accept COVID-19 vaccine (APR=1.13; 95% CI: 1.04, 1.23) compared to construction and Mechanic workers. This finding is consistent with findings from studies done by (1,19) and this could be attributed to ease with access to information among government workers and students as opposed to constructional workers.

### Strengths and limitations

This is the first study conducted in Central Uganda (Wakiso), using an urban based population surveillance cohort data with bigger sample size to report on willingness to receive to COVID-19 vaccine, associated factors and reasons for hesitancy or refusal among persons aged 13-80 years.

Although the cross-sectional nature of this study was the ideal method since both the dependent and independent variables could be measured at the same time. This type of design cannot assess the temporal relationship and therefore more advanced designs could close this gap. Additionally, this study being purely quantitative, it could not explore beliefs, attitudes and perceptions about COVID-19 vaccine hesitancy, hence future qualitative studies are recommended.

## Conclusion

In this study, over 30% of the population were hesitant to accept the COVID-19 vaccine. Young age groups were less likely to receive COVID-19 vaccine while post-primary level of education, and government workers were associated with greater willingness to take the COVID-19 vaccine. The Willingness to accept the COVID-19 vaccine is still low compared to WHO recommended rate. Therefore, efforts to address some of the hesitation reasons like concerns about side effects of the vaccine, myths and misconceptions like vaccines are not effective, will not get seriously ill, getting corona virus from the vaccine.

## Data Availability

Data supporting the conclusions of this article can be accessed in the supporting information labelled as dataset and COVID 19 module_q'nnaire_Eng

## Authors’ contributions

AD contributed to the conception of this research idea, study design, data analysis and including supporting all stages of this paper. RN, AK, GKN, FN, RB, EK, GN, GK, JK, and SM supported with the study design, study conceptualization, data analysis, and interpretation of findings.

## Acknowledgements

We would like to thank Population health surveillance (PHS) participants for their participation in the study.

## Disclaimer

The findings and conclusions in this report are those of the author(s) and do not necessarily represent the official position of Africa Medical & Behavioral Sciences Organization (AMBSO).

